# Evaluation of the awareness level of Healthcare workers toward NCOVID-2019 in Pakistan

**DOI:** 10.1101/2020.03.26.20044636

**Authors:** Bushra Imdad, Uzair Abbas, Ambrina Qureshi, Sehrish Mohsin, Amna Shireen, Altaf Hussain, Ramsha Ali Baloch, Niaz Hussain, Nazia Imdad

## Abstract

**Background:** Novel Coronavirus infection disease 2019 (NCOVID-19), caused by the corona virus, was first spotted in Wuhan, city of China, December 2019. The NCOVID-19 virus is spread among individuals through close communication in the form of droplets, not via airborne. Those individuals are at risk of infection who are in close contact with a NCOVID-19 patient or who take care of NCOVID-19 patients. Infection prevention and control measures are critical to prevent the possible spread of any infection in healthcare facilities. Therefore, healthcare workers should be aware of basic knowledge and all procedures concerning prevention and protection from NCOVID-19.

**Objective:** The objective of this study was to evaluate the awareness level of healthcare workers toward NCOVID-2019 in Pakistan.

**Material and Methods:** A questionnaire was generated according to WHO information that was circulated among the healthcare workers of different hospitals and medical institutes of Pakistan. Calculated sample size was 650.

**Conclusion:** Healthcare workers have insufficient knowledge of preventive measures and infection control. The authorities must take initiatives on urgent basis to increase the awareness among the healthcare workers and general public also so that the drastic circumstances can be avoided in the developing country like Pakistan.

## Introduction

The recognition of Human coronaviruses (HCoV) firstly came during the 1960s in the noses of patients with the nasopharyngitis. There are some known species of coronavirus 229E, NL63, OC43, and HKU1 in which two species of coronaviruses, OC43 and 229E are responsible to affect a huge percentage of common colds.(1) In Latin “Corona” means” crown. The name of Coronaviruses was given due to the crown-like projections on their surfaces. Infections are frequently occurring during winter season moreover in early spring because the temperature is favorable for viruses. Therefore, this is very common an individual easily get infected with coronavirus. An antibody of coronavirus does not last for a long time. Additionally, one strain of the antibodies of coronavirus may be useless against other strains.(2)

Novel corona viruses are those viruses that usually cause pathogenesis in mammals along with human beings by affecting their respiratory tract. These viruses are linked with the conditions like Severe acute respiratory syndrome (SARS), common cold and pneumonia. Additionally, they are also capable of affecting the gut. In 1937 coronavirus was first isolated from an infectious bronchitis virus in birds that could significantly ravage poultry stocks. Almost 15 and 30 percent of common cold are caused by these viruses. During the last 70 years, scientists revealed that coronaviruses are able to contaminate mice, rats, horses, pigs, and cattle, dogs, cats, turkeys.(3)

Different species of human coronaviruses includes 229E, NL63, OC43, and HKU1, typically source of mild to moderate upper-RTI, like the common cold. It is high in chance that people can get affected or fell ill by these viruses at some point in their life. These infections mainly last only for a specific period. Runny nose, headache, cough, sore throat, fever are included in coronaviruses symptoms. Human coronaviruses can at times cause lower-respiratory tract sicknesses, like pneumonia or bronchitis. People with cardiopulmonary disease, Compromised immunity, children and older adults are at more risk.(4, 5) Different studies have been reported two more types of human coronaviruses, MERS-CoV and SARS-CoV are responsible to cause severe respiratory symptoms. MERS have symptoms like cough, fever and shortness of breath which often lead to pneumonia. The reported patients with MERS are around 3/10 and they have died.(3, 6, 7) SARS symptoms comprised of having fever, chills, and body aches which usually progressed to pneumonia. Since 2004 there is no human case reported in the world with SARS.(8) This study was contained the questioner-based knowledge of healthcare workers regarding the current pandemic outbreak of novel corona virus-19.

Novel Coronavirus disease 2019 (NCOVID-19), caused by the corona virus, was first spotted in Wuhan, city of China, in the month of December 2019. (9) The Director-General of WHO has stated on 30 January 2020, that the current outbreak commenced a health emergency of worldwide concern. On account of available evidence, the NCOVID-19 virus is spread among individuals through close communication in the form of droplets, not via airborne. Those individuals are at risk of infection who are in close contact with a NCOVID-19 patient or who take care of NCOVID-19 patients especially healthcare workers. This is very critical to prevent the possible spread of any infection in healthcare facilities. Therefore, healthcare workers should be aware of basic knowledge and all procedures concerning prevention of and protection from NCOVID-19.(10)

### Route of Transmission

Novel coronaviruses can spread through direct or indirect routes. Such as, one person infected with virus infected other non-infected person via air by sneeze, cough and close personal interaction(11), like shake-hands, touch an object or surfaces contaminated with coronavirus, and touch other body part specifically mouth, nose or eyes.(12, 13) Infrequently, spread through fecal route. Coronavirus survive in the fall and winter season therefore; breakout with common human coronaviruses in this season is on large scale. Though, people become infected at any time of the year. Recently it has been reported that in Germany one case of 2019-nCoV infection shows that the virus may also spread by contact with patients without symptoms.(14)

### Prevention and Treatment

Presently, there are no medications and vaccination are available to protect against novel coronavirus infection. Coronavirus can live 12 hours on metal surfaces and 9 hours on fabric. The only way to protect from this virus is to Wash hands with soap and water again and again. Frequently use a hand sanitizer alcohol based and prevent touching the body parts like eyes, nose, and mouth with unwashed and dirty hands. Face to face or close contact with others should be avoided, Mouth and nose should be covered with a tissue when you sneeze or cough, used tissue should be thrown in the dustbin and hands should be washed. Clean and disinfect objects and surfaces.(9, 15)

## Materials and Methods

### Study Population

The study participants were recruited from the healthcare workers of basic health units and primary, secondary and tertiary healthcare facilities across the major cities of Pakistan from the province of Sindh, Punjab, Baluchistan and Gilgit Baltistan. The study also included the undergraduate students who are actively involved in clinical rotation and are in direct contact with patients. The questionnaire containing 16 questions was formulated based on the information given by WHO for NCOVID-19.

The questionnaire was divided into three sections in which 19 multiple questions were asked to assess the knowledge, source of infection, symptoms, protection, infection control, and treatment a single-item scale was used to record the respondents’ replies. Taking 1 million health workers in Pakistan, with 97% confidence interval, the sample size calculated was 650 using OpenEpi. Ethical approval from the institute could not be sought due to the prevailing emergency situation/ lockdown in the country institutions; however, consent from each participant was sought by they filled the online questionnaire. The results were analyzed on SPSS version 24.0. The duration of the study was 10 days.

### Inclusion Criteria

Medical doctors, volunteers and paramedical staff working at different healthcare facilities along with undergraduate medical and dental students who were in direct contact with the patients during their clinical rotations were included in the study.

## Results

This study was based on online questionnaire, which was divided into multiple sections to assess knowledge, source of infection, symptoms, protection, infection control, and treatment. Total 653 participants were recruited in this study from different hospital and medical institutes across Pakistan. The participants comprised 208 (31.85%) doctors, 347 (53.14%) paramedical staff and 98 (15.01%) undergraduate students who are voluntarily involved in clinical rotation in different hospitals of Pakistan.

Out of total, 65.69% of participants opted the correct definition of NCOVID-19 and 28.60% were unaware of the correct definition of NCOVID-19, while 5.72% of participants had no idea. More than one-quarter (77.47%) of the total participants answered that NCOVID-19 is a pandemic outbreak whereas, 10.53% replied that they did not know, 9.40% responded epidemic and 2.59% endemic. Moreover, 57.97% of participants had attained their information about NCOVID-19 from media, 20.32% from social community and 3.06% from college, whereas 0.65% of participants had no idea. Most of the participants 86.99% considered that NCOVID-19 is contagious while, 3.58% participants replied that NCOVID-19 is not contagious and 9.43% has no idea. Almost all of the participants (97.09%) answered NCOVID-19 spreads from infected person with close contact while 1.62% answered contact with infected animal, 0.81% fast food, 0.48% had no idea. Additionally, 37.54% of respondents opted that NCOVID-19 is same as SARS and MERS whereas, 30.15% replied “not same” and 32.31% answered no idea. (Figure-1)

**Figure-1:**
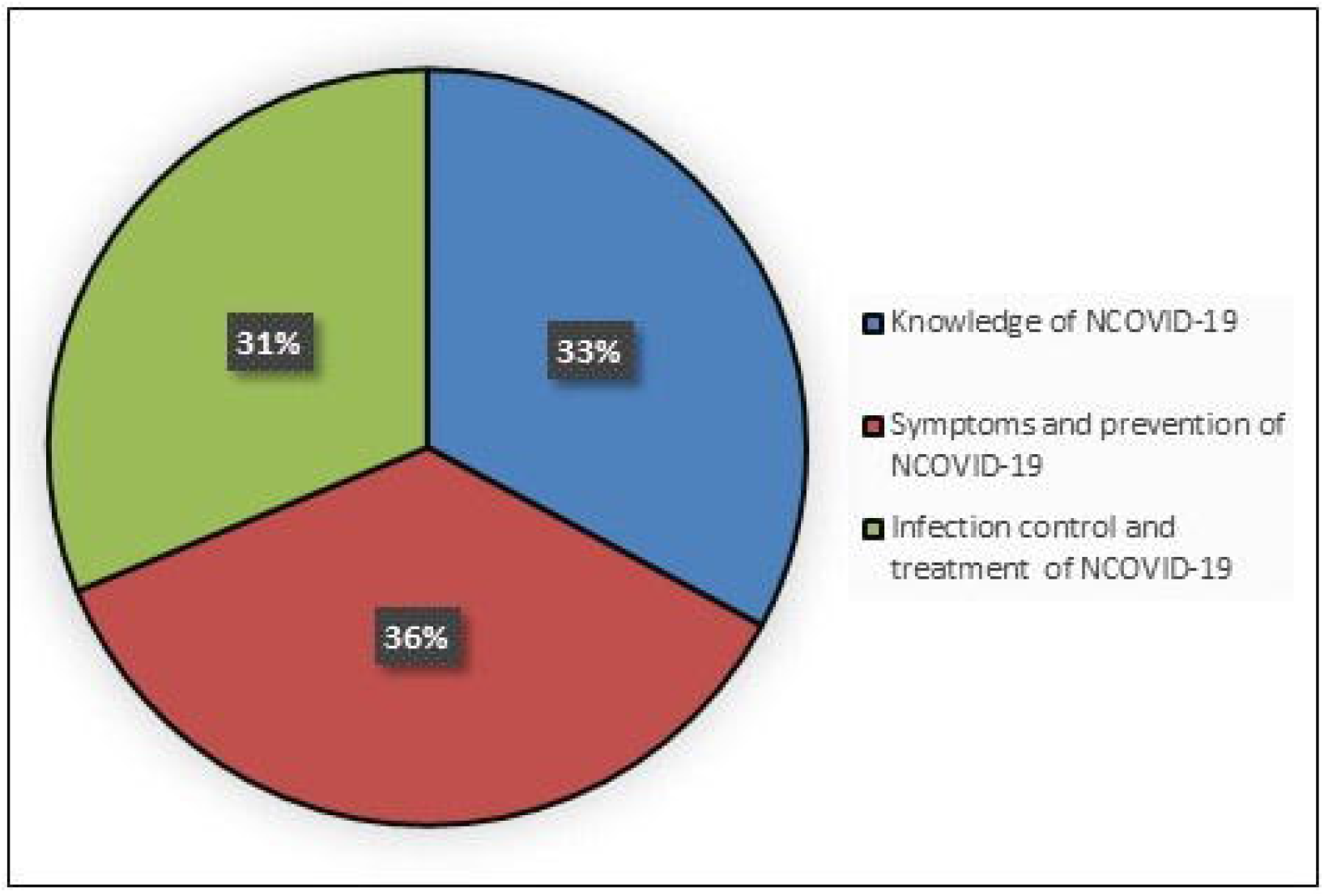
This figure shows that 6 questions related to knowledge about NCOVID-19 where 64.76% participants chose correct option of QNO-1. While, 77.61% participants selected correct option of QNO-2, 75.88% participants chosen correct option of QNO-3, 87.23% participants selected correct option of QNO-4, 96.94% participants selected correct option of QNO-5 and 37.54% participants selected correct option of QNO-6

Furthermore, 95.80% respondents knew related symptoms such as difficulty in breathing, fever and cough whereas, 2.10% participants responded only difficulty in breathing, 0.81% participants answered only fever, 0.65% participants replied only cough and 0.65% replied no idea. Huge percentage (92.41%) of the participants were aware of the preventive measures for NCOVID-19. More than half of the participants (69.53%) answered that the surgical mask can protect from the disease. (Figure-2)

**Figure-2:**
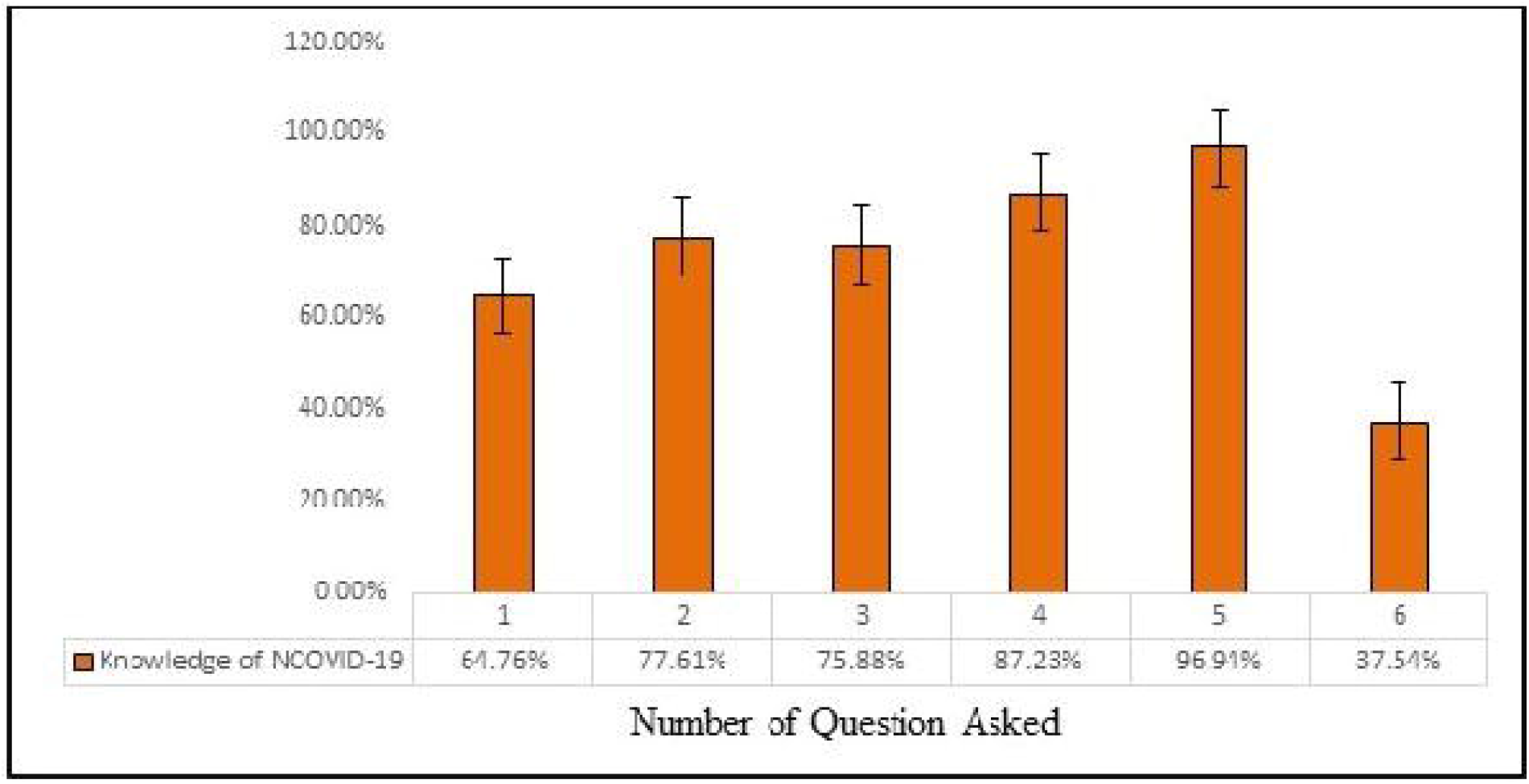
This figure shows 4 questions related to symptoms and preventive measures of NCOVID-19 where 95.72% of participants opted correctly for QNO-1. While, 92.66% participants selected correct option of QNO-2, 69.79% participants selected correct option of QNO-3 and 63.40% participants selected correct option of QNO-4.

More than two third 83.84% of the respondents were well informed that the corona virus can live on surfaces and 48.40% of participants knew the period of incubation for corona virus is few hours up to several days. More than one-third of the participants stated that the rate of mortality of NCOVID-19 is about 2% globally and 66.13% knew that there is no treatment available for NCOVID-19. Additionally, 92.38% of the participants were aware that the effected patients recoverable from the disease and 24.64% opted that the people recovered from the disease can still transfer or spread it whereas, 46.19% answered “NO” and 29.17% replied No idea. Majority of the participants 94.49% answered that the infected person should be isolated and 63.92% of the participants chosen that the isolation ward is available in their setup while, 21.68% replied “NO” and 14.40% had no idea. More than two third of the participants 80.45% answered that elders > 60 years are at higher risk while, 9.53% of the participants replied adults, 6.14% of the participants answered infants and 3.88% of the participants answered newborns. (Figure-3)

**Figure-3:**
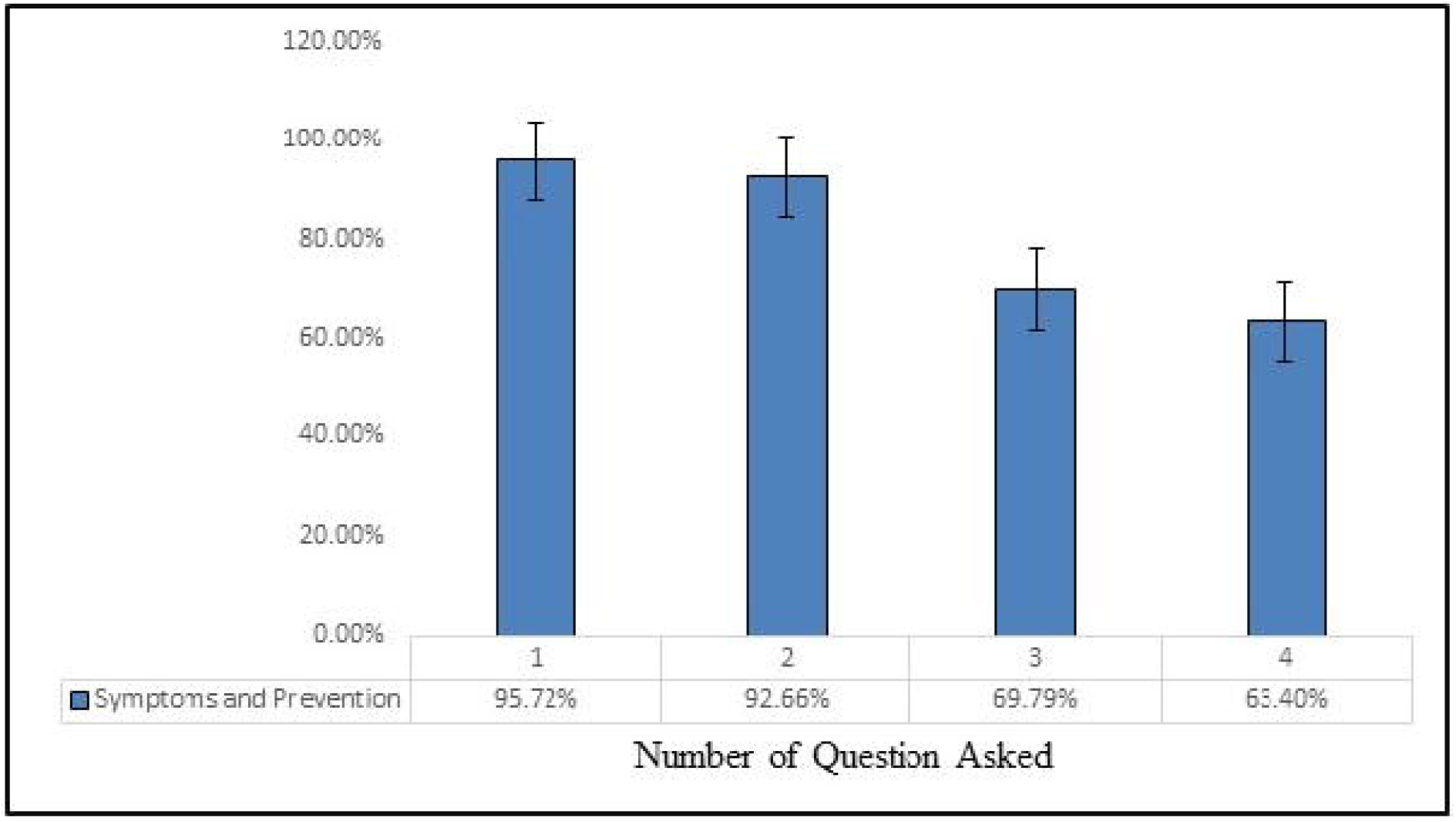
The figure shows 9 questions related to infection control and treatment of NCOVID-19 where 82.87% of participants selected correct option of QNO-1. While, 47.85% participants selected correct option of QNO-2, 63.19% participants chosen correct option of QNO-3, 65.80% participants selected correct option of QNO-4, 92.48% participants selected correct option of QNO-5, 46.93% participants selected correct option of QNO-6, 54.69% participants chosen correct option of QNO-7, 93.56% participants selected correct option of QNO-8, 80.43% participants selected correct option of QNO-9.

## Discussion

Since December 2019, the recently discovered novel coronavirus (2019-nCov) has caused the outbreak in Wuhan the city of China.(16) Now, it has converted into pandemic outbreak. The rapidly increasing number of cases and evidence of human-to-human transmission suggested that the virus was more contagious than SARS-CoV and MERS-CoV.(17-19) a large number of infections of healthcare workers have been reported and the specific reasons for the failure of protection need to be further investigated.(19). Reported in recent study that >3000 (including clinical diagnosis) healthcare workers have been infected with NCOVID-19.(20) Therefore, it is need of the hour that healthcare worker should have proper information about knowledge, prevention, infection control measures and available treatment of options of current pandemic to control its further spread because healthcare workers are the first line combaters of the outbreak and the second line transmission of disease. According to WHO report-60 on 19^th^, March 2020 there are 302 persons infected in Pakistan with the Corona virus and on an average 61 new cases are being reported every day. However, this is not confirmed up till now that how many healthcare workers are confirmed or suspected with infection.

As this study was based on questionnaire, which was divided into three main sections including knowledge, source of infection, symptoms, protection, infection control, and treatment, our study indicated that the 33 % of participants had enough awareness about the definition and source of infection of NCOVID-19. Moreover, 36% participants were aware about the symptoms and how to take preventive measures for NCOVID-19 and 31% of participants had awareness of infection control and treatment of NCOVID-19 (Figure-4). Our data showed that there is insufficient awareness of healthcare workers about NCOVID-19 here in Pakistan.

**Figure-4:**
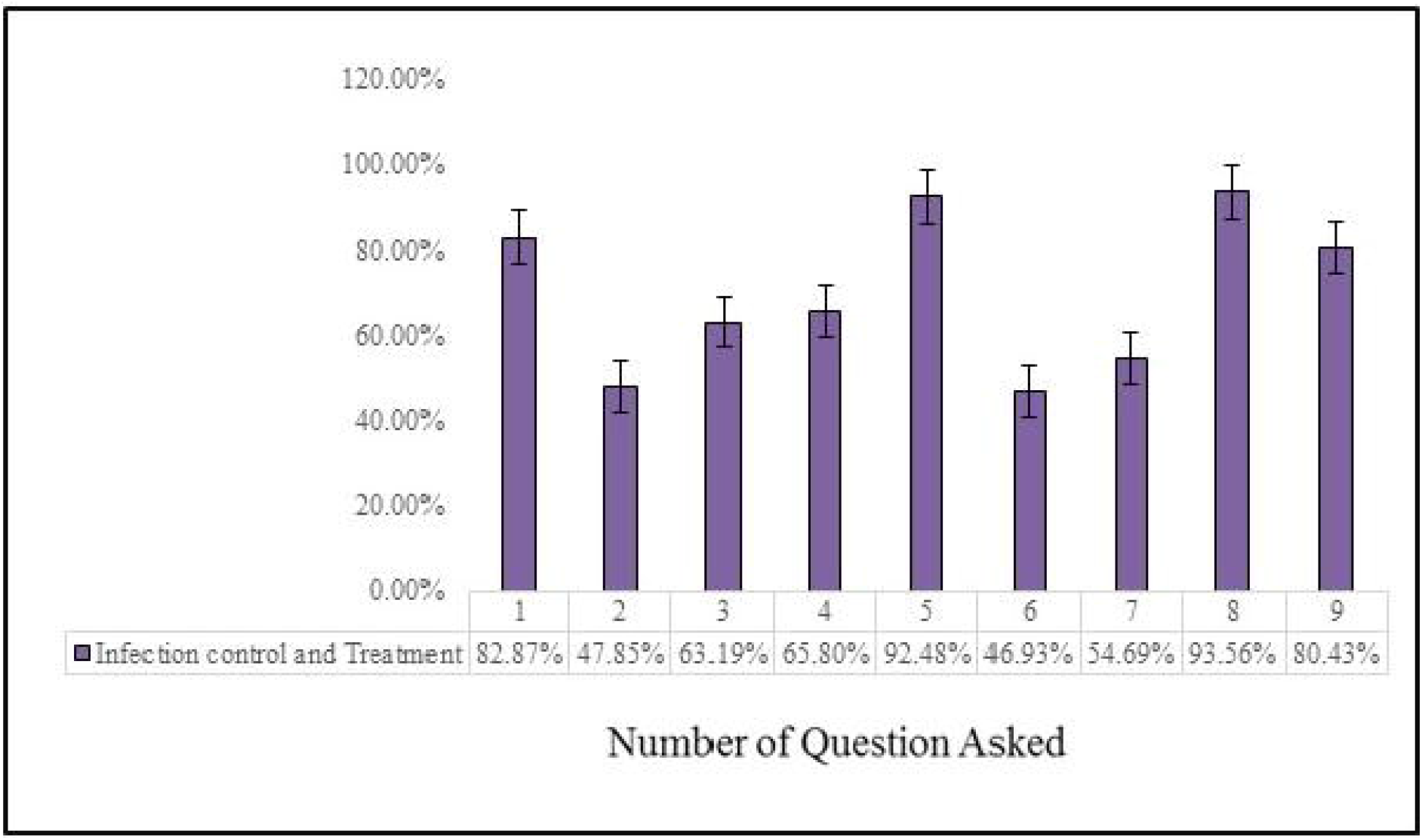
Summarized assessment of the study

Our data shows the requirement for further continuing awareness education among healthcare workers as well as improving public awareness about infection control. The pandemic outbreak of NCOVID-19 and the increasing number of affected people day by day, there is an immense need to acquire the basics information of infection control which is the main principle for protection from NCOVID-19.

## Conclusion

The important way to avoid the disease is to have proper knowledge and preventive measures. Healthcare workers have insufficient knowledge of preventive measures and infection control. The authorities must take initiatives on urgent basis to increase the awareness among the healthcare workers and general public also so that the drastic circumstances can be avoided in the developing country like Pakistan. This data has given us a clue about how serious our healthcare workers are in dealing with this pandemic issue.

## Data Availability

Yes - all data are fully available without restriction

